# Diagnostic accuracy of swab-based molecular tests for tuberculosis using novel near point-of-care platforms: A multi-country evaluation

**DOI:** 10.1101/2025.04.12.25325603

**Authors:** Amy Steadman, Kingsley Manoj Kumar, Lucy Asege, Midori Kato-Maeda, Job Mukwatamundu, Kinari Shah, Trinh Trang, Alexey Ball, Khushboo Khimani, Dao Thi Kim Dung, Joy Sarojini Michael, Devasahayam J Christopher, Ha Phan, Seda Yerlikaya, Payam Nahid, Claudia M. Denkinger, Adithya Cattamanchi, Alfred Andama

## Abstract

**Background:** Swab-based molecular platforms that enable testing of both sputum (via swabs swirled in sputum) and tongue swabs are emerging as a promising option for more accessible and lower cost molecular testing for tuberculosis (TB). We conducted a multi-country evaluation of two novel swab-based molecular tests: Molbio Truenat MTB Ultima (MTB Ultima) and Pluslife MiniDock MTB Test (MiniDock MTB).

**Methods:** Consecutive people ≥12 years old with presumptive TB were enrolled at outpatient health centers in India, Uganda, and Vietnam. We collected two tongue swabs and prepared two sputum swabs for MTB Ultima and MiniDock MTB testing, then evaluated the diagnostic accuracy of MTB Ultima and MiniDock MTB using both swab types against a sputum liquid culture-based microbiological reference standard (MRS). The diagnostic accuracy of swab-based molecular tests was also compared to sputum Xpert MTB/RIF Ultra (Xpert Ultra) and auramine smear microscopy.

**Findings:** From January to September 2024, 1,050 participants were included in the tongue swab MTB Ultima evaluation, 197 in the sputum swab MTB Ultima evaluation, and 322 in the MiniDock MTB evaluations. In comparison to sputum Xpert Ultra, sensitivity was similar for both sputum swab MTB Ultima (93.6% vs. 100.0%, difference -6.4%, [95% CI: -15.5, 2.7], p=0.25) and MiniDock MTB (91.0% vs. 94.0%, difference –3.0% [95% CI: -8.6, 2.6], p=0.50). In comparison to sputum smear microscopy, sensitivity was higher for both tongue swab MTB Ultima (77.9% vs. 59.1%, difference 18.8% [95% CI: 10.8, 26.8], p<0.0001) and MiniDock MTB (85.7% vs. 67.1%, difference 18.6% [95% CI: 7.2, 29.9], p=0.001). Specificity was high (>98%) for both tests with sputum swabs and tongue swabs.

**Interpretation:** MTB Ultima and MiniDock MTB have similar accuracy to current sputum-based molecular tests with sputum swabs and meet minimum accuracy thresholds for a non-sputum, near point-of-care molecular test with tongue swabs. These tests offer strong potential to make universal molecular testing for TB a reality.

## Introduction

Missed or delayed diagnosis of tuberculosis (TB) remains common despite the availability and scale-up of World Health Organization (WHO)-recommended rapid molecular tests since 2010.^1– 4^ A key reason is that current molecular tests cannot be deployed at lower-level health facilities in high burden countries due to cost, infrastructure and/or human resource requirements.^5^ WHO-recommended rapid diagnostics were available in less than half of TB diagnostic units in 23 of 30 high burden countries and were used as the initial test for TB in less than half of people with TB in 2024. As a result, the ongoing failure to rapidly diagnose and treat TB leads to excess morbidity, mortality, and disease transmission.^1^

Leveraging advances made during the COVID-19 pandemic, novel swab-based platforms are emerging that respond to the urgent need for simpler molecular tests for TB. In contrast to whole-sputum-based testing, swab-based testing avoids the need for homogenization, nucleic acid extraction and purification, thereby reducing cost and complexity.^6,7^ Two swab-based tests are at advanced stages of development. Truenat MTB Ultima (MTB Ultima; Molbio Diagnostics, India) replaces DNA extraction with a simple sonication-based automated mechanical lysis step (Truelyse) followed by DNA amplification using an existing automated real-time quantitative microPCR analyzer (Truelab). Similarly, MiniDock MTB (Guangzhou Pluslife Biotech, China) employs an automated heat-based mechanical lysis step (Thermolyse) followed by isothermal amplification using a novel method originally developed for SARS-CoV-2 testing.^8^ Both platforms are battery-operated and have simple workflows (**Figure 1**), making them amenable for use by health workers without laboratory training in peripheral health facilities. Further, both platforms enable testing of sputum (via swabs swirled in sputum) and tongue swabs, which are emerging as an alternative sample type for TB detection.^9–20^

**Figure 1.**
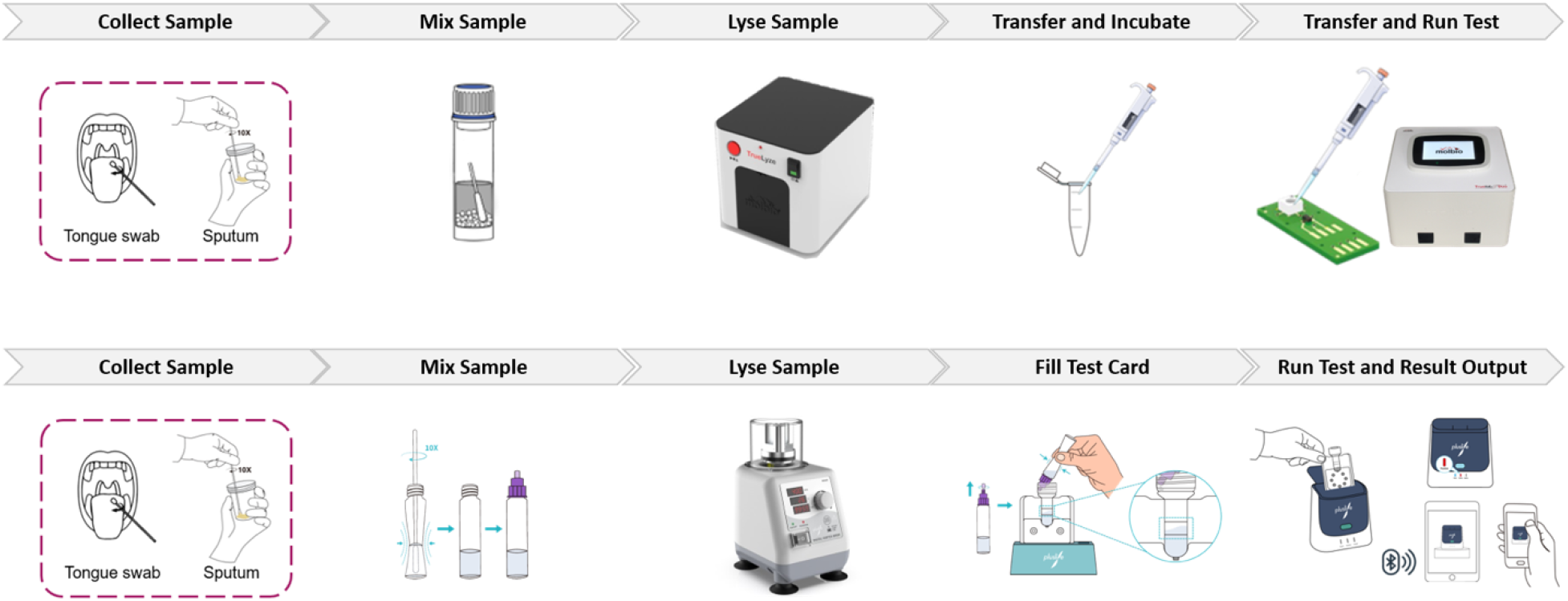
Operating procedures for MTB Ultima (top panel) and MiniDock MTB (bottom panel).

We conducted a multi-country, prospective evaluation of the diagnostic accuracy of late-prototype versions of MTB Ultima and MiniDock MTB against a microbiological reference standard (MRS), comparing their performance to current sputum-based tests (smear microscopy and Xpert MTB/RIF Ultra [Xpert]) and to the WHO-recommended minimum accuracy targets for a near point-of-care (POC) TB diagnostic test.^21^

## Methods

### Study design and population

We conducted a prospective, cross-sectional, multi-center diagnostic accuracy study at outpatient health centers in India, Uganda, and Vietnam as part of the Rapid Research in Diagnostics Development for TB Network (R2D2 TB Network, ClinicalTrials.gov NCT04923958).^21,22^ We consecutively enrolled non-hospitalized adolescents and adults aged 12 years or older who had presumptive TB based on: 1) a new or worsening cough lasting ≥2 weeks, or 2) a risk factor for TB (HIV infection, self-reported close contact with a TB patient, or a history of mining work) plus an abnormal WHO-recommended TB screening test (abnormal CXR or, for people living with HIV [PLHIV], C-reactive protein [CRP] >5 mg/L). We excluded individuals who had been treated for TB infection or disease in the past 12 months, had taken antibiotics with antimycobacterial activity in the two weeks prior to enrolment, or were unable (residing >20 km from the study site) or unwilling to return for follow-up or provide informed consent. Based on availability of the applicable supplies and standard operating procedures for each test and specimen type, enrolment occurred between January and September 2024 for the tongue swab MTB Ultima evaluation, May and September 2024 for the sputum swab MTB Ultima evaluation, and between April and September 2024 for both the tongue and sputum swab MiniDock MTB evaluations. This is an initial validation study aimed at estimating the diagnostic accuracy of the novel test. The study was designed and reported in accordance with the Standards for Reporting of Diagnostic Accuracy Studies (STARD) guidelines.^23^

### Procedures

Eligible participants completed a demographic and clinical questionnaire, including TB history. Participants were asked to provide finger prick or venous blood, up to two tongue swabs, and up to three spot sputum samples (including by sputum induction for participants unable to expectorate). Tongue swabs were collected prior to sputum collection by swabbing the dorsum of the tongue for 30 seconds with a nylon flocked swab (Copan 520CS01 for MTB Ultima and Copan 502CS01 for MiniDock MTB) as described previously.^24^ The first sputum sample was used to generate two sputum swabs by swirling a nylon flocked swab (Copan 520CS01 for MTB Ultima and Copan 502CS01 for MiniDock MTB) 10 times for 15 seconds and then wiping the swab against the inside wall of the sputum container. After preparation of sputum swabs, the remainder of the first sputum sample was used for Xpert testing. The second and third sputum samples were decontaminated with N-acetyl-L-cysteine/sodium hydroxide and tested with liquid culture (MGIT 960, BD Microbiology Systems) x 2 and LED fluorescence microscopy with Auramine staining x 2, according to manufacturer or standard guidelines.^25–27^ Xpert Ultra testing was repeated if the first result was invalid or returned a “Trace” semiquantitative value. Sputum Xpert, smear, and culture reference testing were performed by laboratory personnel blinded to the results of all novel index tests.

### Index tests

Tongue swabs and sputum swabs were immediately placed into manufacturer-supplied tubes prefilled with proprietary buffer and tested with MTB Ultima or MiniDock MTB within 24 hours in accordance with manufacturer instructions (see **Supplemental Methods** for details). MTB Ultima results were classified as positive (“MTB Detected”), negative (“MTB Not Detected”) or invalid (*i*.*e*., the endogenous control was not detected). Positive results were further characterized semi-quantitatively as Very Low, Low, Medium or High based on manufacturer-defined Cycle threshold (Ct) ranges. MiniDock MTB results were classified as positive, negative, or invalid. Swab testing was done in a blinded manner (*i*.*e*., before results of sputum tests were known), and was repeated once using leftover sample if the initial result was invalid.

### Reference Standard

We used a microbiological reference standard (MRS) based on culture results. Participants were considered positive for TB if either of the two liquid cultures were positive for *Mycobacterium tuberculosis* (MTB). Participants were considered negative for TB if both cultures were negative. Participants were considered to have indeterminate TB status and excluded from the analysis if no cultures were positive but one or both cultures were contaminated.

### Sample size

The overall sample size was based on the tongue swab MTB Ultima evaluation. Sample size was calculated using the Buderer method to estimate precision around a target sensitivity estimate compared to the MRS.^28^ Assuming a target sensitivity of 75% and disease prevalence of 15%, a minimum of 129 TB-positive participants were required to achieve ±7.5% precision.

### Data analysis

We calculated sensitivity and specificity of tongue swab- and sputum swab-based index tests (MTB Ultima and MiniDock MTB) and sputum-based comparator tests (smear microscopy and Xpert Ultra) along with their exact binomial 95% confidence intervals overall and in key sub-groups (country, sex, diabetes status, HIV status). We compared sensitivity and specificity differences between index and comparator tests using McNemar’s test of paired proportions. Primary analyses considered only the first index test result. For comparator tests, sputum smears were classified as positive when ≥1 AFB was detected per 100 fields. Xpert Ultra results were classified as positive for positive results with a semi-quantitative grade of “Very Low” or higher. We conducted pre-specified sensitivity analyses that included repeat index test results and that counted sputum Xpert Ultra “Trace” results as positive. Secondarily, we assessed concordance of swab-based index tests with sputum Xpert Ultra results overall and by Xpert Ultra semi-quantitative grade.

### Ethics

Ethical approval was granted by the institutional review boards or research ethics committees at the University of California San Francisco (USA; 20-32670), University of Heidelberg (Germany; S-539/2020), Makerere University (Uganda; 2020-182), Christian Medical College (India; 13256), and National Lung Hospital (Vietnam; 22/BVPHN) (see **Supplemental Methods**). Written informed consent was obtained from all participants. The study is registered with ClinicalTrials.gov (NCT04923958).^22^ All study data were collected and managed in a secure Research Electronic Data Capture (REDCap) database hosted at the University of California San Francisco.

## Results

Between January and September 2024, we enrolled 1,127 participants meeting the eligibility criteria (**Figure 2**). Median age was 44.7 years (IQR 30-59), 46.4% were female, 11.1% were people living with HIV (PLHIV), 22.2% were people living with diabetes (PLWD), and 14.8% had confirmed TB based on the MRS (**Table 1**). Demographic and clinical characteristics were similar among subsets of participants included in each index test evaluation (**Supplemental Tables S1-S2**).

**Table 1.** Demographic and clinical characteristics.

|  | Overall<br>N=1,127 | India<br>N=440 | Uganda<br>N=407 | Vietnam<br>N=280 |
| --- | --- | --- | --- | --- |
| Age (median, IQR) | 44.7 (30.0-59.0) | 50.5 (39.5-63.0) | 34.1 (24.0-43.0) | 51.1 (37.0-65.0) |
| Female | 523 (46.4%) | 181 (41.1%) | 211 (51.8%) | 131 (46.8%) |
| Person living with HIV | 124 (11.1%) | 4 (0.9) | 119 (29.2%) | 1 (0.4%) |
| Person living with diabetes | 250 (22.2%) | 125 (28.4%) | 85 (20.9%) | 40 (14.3%) |
| Prior TB | 129 (11.4%) | 44 (10.0%) | 44 (10.8%) | 41 (14.6) |
| Sputum smear-positive | 100 (8.9%) | 22 (5.0%) | 63 (15.5%) | 15 (5.4%) |
| Sputum Xpert Ultra-positive | 152 (13.6%) | 34 (7.8%) | 88 (21.6%) | 30 (10.7%) |
| Very low | 17 (11.2%) | 3 (8.8%) | 7 (7.9%) | 7 (23.3%) |
| Low | 48 (31.6%) | 14 (41.2%) | 24 (27.3%) | 10 (33.3%) |
| Medium | 34 (22.4%) | 8 (23.5%) | 17 (19.3%) | 9 (30.0%) |
| High | 53 (34.9%) | 9 (26.4%) | 40 (45.4%) | 4 (13.3%) |
| Culture (MRS)-positive | 167 (14.8%) | 34.0 (7.8%) | 90 (22.1%) | 42 (15.0%) |

Abbreviations: IQR, inter-quartile range; TB, tuberculosis; MRS, microbiological reference standard.

**Figure 2.**
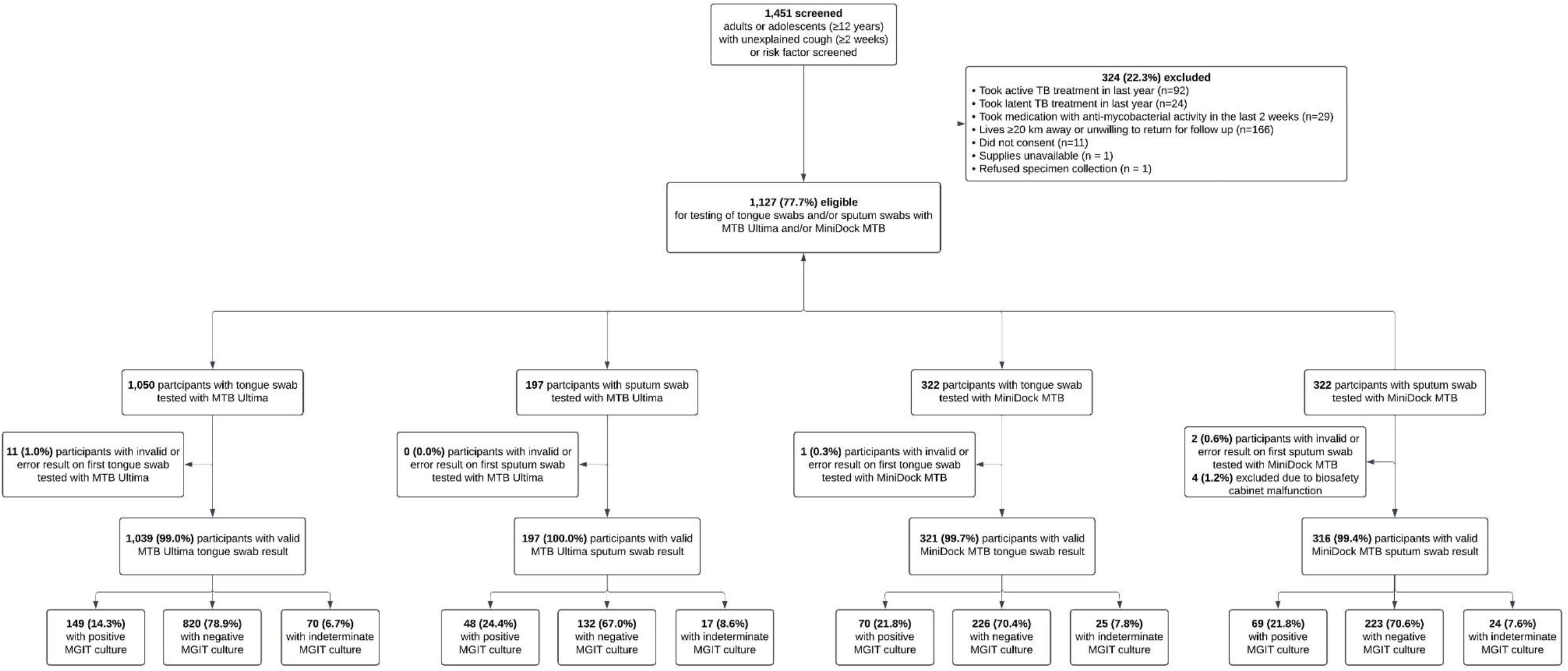
Participant flow diagram.

### Sputum swabs

Sputum swabs were collected from 197 participants for MTB Ultima testing and 322 participants for MiniDock MTB testing. No sputum swab MTB Ultima results and only 2 (0.6%) MiniDock MTB results were invalid; with repeat testing of leftover lysate, all participants had valid results (**Table 2**). Four (1.25%) sputum swabs collected for MiniDock MTB were excluded from testing due to a malfunction of a biosafety cabinet.

**Table 2.** Invalid test results.

|  | **Overall**<br>**n/N (%)** | **India**<br>**n/N (%)** | **Uganda**<br>**n/N (%)** | **Vietnam**<br>**n/N (%)** |
| --- | --- | --- | --- | --- |
| **MTB Ultima** |  |  |  |  |
| Tongue swab initial test | 11/1,050<br>(1.0%) | 4/440<br>(0.9%) | 4/367<br>(1.1%) | 2/243<br>(1.2%) |
| Tongue swab repeat test | 0/7<br>(0.0%) | - | 0/4(0.0%) | 0/3<br>(0.0%) |
| Sputum swab initial test | 0/197<br>(0.0%) | - | 0/169<br>(0.0%) | 0/28<br>(0.0%) |
| Sputum swab repeat test | N/A | - | N/A | N/A |
| **MiniDock MTB** |  |  |  |  |
| Tongue swab initial test | 1/322<br>(0.3%) | - | 0/257<br>(0.0%) | 1/65<br>(1.5%) |
| Tongue swab repeat test | 0/1<br>(0.0%) | - | N/A | 0/1<br>(0.0%) |
| Sputum swab initial test | 2/318<br>(0.6%) | - | 0/253<br>(0.0%) | 2/65<br>(3.1%) |
| Sputum swab repeat test | 0/2<br>(0.0%) | - | N/A | 0/2<br>(0.0%) |
| **Xpert Ultra** |  |  |  |  |
| Sputum initial test* | 3/1,121<br>(0.3%) | 0<br>(0.0%) | 0<br>(0.0%) | 3/280<br>(1.1%) |

\*Unable to obtain sputum samples from six participants in India

Sputum swabs tested with MTB Ultima had high sensitivity (44/48, 91.7% [95% CI: 80.0, 97.7]) and specificity (129/132, 97.7% [95% CI: 93.5, 99.5]) (**Table 3**). When compared to the same sample set, MTB Ultima had higher sensitivity than sputum smear microscopy (91.7% vs. 72.9%, difference 18.7% [95% CI: 5.6, 31.9], p=0.0039) and similar sensitivity to sputum Xpert (93.6% vs. 100.0%, difference -6.4%, [95% CI: -15.5, 2.7], p=0.25).

**Table 3.** Diagnostic accuracy of swab-based MTB Ultima and sputum comparator tests.

|  | **Sensitivity\***<br>**n/N, % (95% CI)** | **Specificity\***<br>**n/N, % (95% CI)** |
| --- | --- | --- |
| **Tongue swab evaluation** |  |  |
| Tongue swab MTB Ultima | 116/149<br>77.9% (70.3, 84.2) | 805/820<br>98.2% (97.0, 99.0) |
| Sputum Xpert Ultra* | 129/139<br>92.8% (87.2, 96.5) | 799/806<br>99.1% (98.2, 99.7) |
| Sputum smear microscopy | 88/149<br>59.1% (50.7, 67.0) | 817/820<br>99.6% (98.9, 99.9) |
| **Sputum swab evaluation** |  |  |
| Sputum swab MTB Ultima | 44/48<br>91.7% (80.0, 97.7) | 129/132<br>97.7% (93.5, 99.5) |
| Sputum Xpert Ultra^ | 47/47<br>100% (92.5, 100.0) | 126/128<br>98.4% (94.5, 99.8) |
| Sputum smear microscopy | 35/48<br>72.9% (58.2, 84.7) | 132/132<br>100% (97.2, 100.0) |

\*Excludes 1 error and 23 trace results on sputum Xpert Ultra
^Excludes 5 trace results on sputum Xpert Ultra

Similarly, sputum swab MiniDock MTB had high sensitivity (62/69, 89.9% [95% CI: 80.2, 95.8]) and specificity (219/223, 98.2% [95% CI: 95.5, 99.3]) (**Table 4**). When compared to the same sample set, MiniDock MTB had higher sensitivity than sputum smear microscopy (89.9% vs. 66.7%, difference 23.2% [95% CI: 11.8, 34.6], p=<0.0001) and similar sensitivity to sputum Xpert (91.0% vs. 94.0%, difference –3.0% [95% CI: -8.6, 2.6], p=0.50).

**Table 4.** Diagnostic accuracy of swab-based MiniDock MTB and sputum comparator tests.

|  | **Sensitivity**<br>**n/N, % (95% CI)** | **Specificity**<br>**n/N (95% CI)** |
| --- | --- | --- |
| Tongue swab MiniDock MTB | 60/70<br>85.7% (75.3, 92.9) | 226/226<br>100% (98.4, 100.0) |
| Sputum swab MiniDock MTB* | 62/69<br>89.9% (80.2, 95.8) | 219/223<br>98.2% (95.5, 99.3) |
| Sputum Xpert Ultra^ | 64/68<br>94.1% (85.6, 98.4) | 220/221<br>99.5% (97.5, 100.0) |
| Sputum smear microscopy | 47/70<br>67.1% (54.9, 77.9) | 226/226<br>100% (98.4, 100.0) |

\*4 participants were not tested with sputum swab MiniDock MTB
^Excludes 2 errors and 5 trace results on sputum Xpert Ultra

Sensitivity of both sputum swab tests was similar across sub-groups defined by sex, HIV status, and diabetes (**Supplemental Tables S3 and S4**). MiniDock MTB higher sensitivity (94.3%, [95% CI: 86.0, 98.4]) and specificity (98.2%, [95% CI: 95.5, 99.5]) when repeat test results were considered (**Supplemental Table S5**). Positive concordance with sputum Xpert results was high across semi-quantitative grades for both sputum swab MTB Ultima (**Supplemental Table S6**) and MiniDock MTB (**Supplemental Table S**7).

### Tongue swabs

Tongue swabs were collected from 1,050 participants for MTB Ultima testing and 322 participants for MiniDock MTB testing. Results were invalid for 11 (1.0%) participants for MTB Ultima and 1 (0.3%) participant for MiniDock MTB. Four tongue swabs with invalid MTB Ultima results were not retested because the remaining lysate was discarded; all other repeat tests returned valid results (**Table 2**).

Tongue swabs tested with MTB Ultima had moderate sensitivity (116/149, 77.9% [95% CI: 70.3, 84.2]) and high specificity (805/820, 98.2% [95% CI: 97.0, 99.0]) (**Table 3**). When compared to the same sample set, MTB Ultima had higher sensitivity than sputum smear microscopy (77.9% vs. 59.1%, difference 18.8% [95% CI: 10.8, 26.8], p=<0.0001) but lower sensitivity than sputum Xpert Ultra (80.6% vs. 92.8%, difference -12.2% [95% CI: -5.4, -19.1], p=0.0002).

Similarly, tongue swabs tested with MiniDock MTB had moderate sensitivity (60/70, 85.7% [95% CI: 75.3, 92.9]) and high specificity (226/226, 100% [95% CI: 98.4, 100.0]). When compared to the same sample set, MiniDock MTB had higher sensitivity than sputum smear microscopy (85.7% vs. 67.1%, difference 18.6% [95% CI: 7.2, 29.9], p=0.001) and lower sensitivity than sputum Xpert (86.8% vs. 94.1%, difference -7.3% [95% CI: -15.0, 0.3] p=0.0625) (**Table 4**).

Sensitivity of both tongue swab tests was similar across sub-groups defined by sex, HIV status, and diabetes. (**Supplemental Tables S8 and S9**). Among participants unable to produce sputum and requiring induction, sensitivity was 71.4% (10/14, [95% CI: 41.9, 91.6]) for tongue swab MTB Ultima and 50.0% (2/4, [95% CI: 6.8, 93.2]) for tongue swab MiniDock MTB; both tests had high specificity in this sub-group (**Supplemental Table S10**).

Sensitivity and specificity were similar for both tests when repeat test results were included (**Supplemental Tables S5 and S11**). Positive concordance with sputum Xpert results was high for the high and medium semi-quantitative categories but progressively lower thereafter for both MTB Ultima (**Supplemental Table S12**) and MiniDock MTB (**Supplemental Table S13**).

## Discussion

We conducted the first multi-country prospective evaluations of two innovative, flexible-specimen, near-POC molecular tests: MTB Ultima and MiniDock MTB. Our results demonstrate that both platforms meet or exceed the WHO TPP requirements for TB diagnostics, with sensitivity and specificity values that surpass or are similar to the minimum thresholds for both sputum-based (>85% sensitivity, >98% specificity) and non-sputum-based (>75% sensitivity, >98% specificity) tests in the near-POC category.^20^ Additionally, both tests exhibited very low failure rates, suggesting that these technologies are robust and suitable for deployment in resource-limited settings where TB diagnostics are urgently needed.

These findings indicate that the MTB Ultima and MiniDock MTB workflows represent a significant advancement over traditional sputum smear microscopy. In particular, both platforms offer enhanced diagnostic accuracy and could serve as viable replacements for sputum smear microscopy in lower-level health facilities. Their efficacy with multiple specimen types, including tongue swabs, introduces a promising alternative to sputum collection—especially when obtaining sputum samples is challenging. This is particularly relevant in populations such as children, PLHIV, elderly persons, and other individuals with difficulty producing sputum.

Furthermore, both systems have the potential to offer cost-effective alternatives to more expensive and/or complex technologies like Xpert and Truenat MTB Plus. MTB Ultima, for example, replaces the Trueprep extraction instrument with the more affordable Truelyse device, reducing the need for consumables. Similarly, MiniDock MTB utilizes the Thermolyse and MiniDock platforms, each priced at approximately $150 USD and a low per test cost (<$4 USD), making it feasible as a replacement for sputum smear microscopy at peripheral health facilities. Both platforms require minimal hands-on time and provide rapid results—less than 45 minutes for MTB Ultima and less than 30 minutes for MiniDock MTB —facilitating timely clinical decision-making and improving patient care.

While the results of this initial evaluation are promising, further validation in larger cohorts and diverse geographical settings is necessary. MiniDock MTB was evaluated in a limited subset of participants, primarily from Uganda, where the majority had higher bacillary loads. Expanding testing to other populations, particularly those with lower bacillary loads or different demographics, will be crucial for understanding the generalizability of the results. Similarly, although the MTB Ultima test was conducted on a limited number of sputum swab specimens, the initial findings indicate that sputum swabs can achieve high sensitivity and low failure rates, even without the use of DNA extraction. Further studies should expand testing of these platforms to a broader range of patient groups and clinical settings, especially patients who cannot expectorate sputum. Notably, the proportions of PLHIV were small in this preliminary evaluation, and studies examining the performance in children <12 years old are ongoing.

Importantly though, the flexibility to test either sputum swabs or tongue swabs provides healthcare workers with a crucial advantage. In situations where sputum is not available, a tongue swab offers a viable alternative while maintaining reasonable sensitivity, especially in comparison to sputum smear microscopy. This flexibility is especially important for populations less likely to expectorate high-quality sputum, such as key populations with higher risk for TB including PLHIV, children, and the elderly. Future studies should assess the diagnostic yield of tongue swabs in these groups who may benefit from non-sputum-based diagnostic options.

Both platforms have the potential to further improve access to TB diagnostics by simplifying testing procedures and reducing the need for specialized laboratory infrastructure. This is a significant advantage in settings where access to centralized laboratories or trained personnel is limited. However, while the initial results are promising, ongoing research and larger-scale studies are needed to fully assess the utility of these platforms in routine clinical practice and to understand their performance in diverse patient populations and geographical regions.

Ultimately, the MTB Ultima and MiniDock MTB tests represent a transformative step forward in tuberculosis diagnostics, offering accessible, flexible, and cost-effective solutions for resource-limited settings. By providing accurate results with minimal infrastructure requirements, these platforms have the potential to revolutionize TB diagnosis and contribute significantly to global efforts in TB control. Further validation and broader implementation studies are essential to maximize their impact and ensure they meet the needs of diverse populations worldwide. With continued research and development, these innovations could become key tools in the fight against tuberculosis, helping to save lives and advance global health equity.

## Data Availability

All data produced are contained in the Supplemental Data file

## Acknowledgements

The authors would like to thank the patients, staff, and administration of Christian Medical College, Hanoi Lung Hospital, Vietnam National Lung Hospital, Mulago National Referral Hospital, and Kisenyi Health Centre for their support and participation in the study.

## Data Sharing

De-identified data may be accessed in the **Supplemental Data** file.

## Author Declarations

Authors confirm all relevant ethical guidelines have been followed, and any necessary IRB and/or ethics committee approvals have been obtained.

