## Supplemental Methods for "Diagnostic accuracy of swab-based molecular tests for tuberculosis using novel near point-of-care platforms: A multi-country evaluation"

**R2D2 enrollment sites and ethics committees**

| City, Country | Enrollment site | Ethics committee |
|---|---|---|
| Vellore, Tamil Nadu, India Chittoor, Andhra Pradesh, India | CMC Pulmonary Outpatient Department, Primary care clinic in Vellore (CHAD) and Chittoor (CMC satellite campus) | Christian Medical College Institutional Review Board (13256) |
| Hanoi, Vietnam | Outpatient departments, Hanoi Lung Hospital | Ministry of Health Ethical Committee for National Biological Medical Research (94/CN-HĐĐĐ); National Lung Hospital Ethical Committee for Biological Medical Research (566/2020/NCKH); Hanoi Lung Hospital Science and Technology Initiative Committee (22/BVPHN) |
| Kampala, Uganda | Mulago Outpatient Department, Kisenyi Health Center | Makerere University, College of Health Sciences, School of Medicine, Research Ethics Committee (2020-182) |

*This study was also approved at the University of California, San Francisco Institutional Review Board (20-32670), and the University of Heidelberg Ethics Committee of the Medical Faculty (S-539/2020).

### Index test methods in detail

| Index test | Methods |
|---|---|
| MTB Ultima | • Swabs placed in tube containing proprietary buffer<br>• 90-second lysis step in the automated Truelyse sonication device to generate lysate<br>• 6µl of lysate added to a freeze-dried reagent tube and incubated for 30-60 seconds<br>• Entire volume transferred to an MTB Ultima chip for amplification in the Truelab PCR platform<br>• Result after 40 minutes |
| Minidock MTB | • Swabs swirled 10 times in tubes prefilled with proprietary buffer<br>• Swab eluate heated and lysed<br>• Tube screw cap removed, tubes inverted to squeeze lysate onto reaction cards up to the volume indicated by the fill line.<br>• Reaction card incubated at ambient temperature for 15 seconds to rehydrate the reagents<br>• Air bag on reaction card screw cap firmly pressed and the reaction cards shaken up and down 10 times in about 5 seconds<br>• Reaction card placed in the MiniDock Pro device for isothermal amplification<br>• Result after 25 minutes |
