## Supplemental Tables for "Diagnostic accuracy of swab-based molecular tests for tuberculosis using novel near point-of-care platforms: A multi-country evaluation"

**Table S1. Baseline characteristics of MTB Ultima on sputum-dipped swab.**

| | Overall<br>N=197 | Uganda<br>N=169 | Vietnam<br>N=28 |
|---|---|---|---|
| Age (median, IQR) | 36.1 (25.0-45.0) | 33.1 (24.0-41.0) | 53.9 (40.0-64.5) |
| Female | 106 (53.8%) | 91 (53.8%) | 15 (53.6%) |
| Person living with HIV | 37 (18.8%) | 37 (21.9%) | 0 (0.0%) |
| Person living with diabetes | 32 (16.2%) | 28 (16.6%) | 4 (14.3%) |
| Prior TB | 18 (9.1%) | 14 (8.3%) | 4 (14.3%) |
| Sputum smear-positive | 35 (17.8%) | 33 (19.5%) | 2 (7.1%) |
| Sputum Xpert Ultra-positive | 49 (24.9%) | 45 (26.6%) | 4 (14.3%) |
| Very low | 3 (6.1%) | 3 (6.7%) | 0 (0.0%) |
| Low | 15 (30.6%) | 13 (28.9%) | 2 (50.0%) |
| Medium | 13 (26.5%) | 11 (24.4%) | 2 (50.0%) |
| High | 18 (36.7%) | 18 (40.0%) | 0 (0.0%) |
| Culture (MRS)-positive | 48 (24.4%) | 44 (26.0%) | 4 (14.3%) |

Abbreviations: IQR, inter-quartile range; TB, tuberculosis; MRS, microbiological reference standard.

**Table S2. Baseline characteristics of MiniDock MTB population**

| | Overall<br>N=322 | Uganda<br>N=257 | Vietnam<br>N=65 |
|---|---|---|---|
| Age (median, IQR) | 36.0 (25.0-45.0) | 32.3 (24.0-40.0) | 50.4 (35.0-64.0) |
| Female | 158 (49.1%) | 126 (49.0%) | 32 (49.2%) |
| Person living with HIV | 63.0 (19.6%) | 63.0 (24.5%) | 0 (0.0%) |
| Person living with diabetes | 57 (17.7%) | 49 (19.1%) | 8 (12.3%) |
| Prior TB | 37 (11.5%) | 24 (9.3%) | 13 (20.0%) |
| Sputum smear-positive | 47 (14.6%) | 45 (17.5%) | 2 (3.1%) |
| Sputum Xpert Ultra-positive | 66 (20.5%) | 60 (23.3%) | 6 (9.2%) |
| Very low | 4 (6.1%) | 3 (5.0%) | 1 (16.7%) |
| Low | 20 (30.3%) | 17 (28.3%) | 3 (50.0%) |
| Medium | 17 (25.7%) | 15 (25.0%) | 2 (33.3%) |
| High | 25 (37.9%) | 25 (41.7%) | 0 (0.0%) |
| Culture (MRS)-positive | 71 (22.0%) | 63 (24.5%) | 8 (12.3%) |

Abbreviations: IQR, inter-quartile range; TB, tuberculosis; MRS, microbiological reference standard.

**Table S3. Diagnostic accuracy of sputum swabs tested with MTB Ultima compared to MRS by subgroups.**

| | **Sensitivity**<br>**n/N, % (95% CI)** | **Specificity**<br>**n/N (95% CI)** |
|---|---|---|
| Overall | 44/48, 91.7 (80.0-97.7) | 129/132, 97.7 (93.5-99.5) |
| Country | | |
| Uganda | 41/44, 93.2 (81.3-98.6) | 106/109, 97.2 (92.2-99.4) |
| Vietnam | 3/4, 75.0 (19.4-99.4) | 23/23, 100.0 (85.2-100.0) |
| Sex | | |
| Female | 18/18, 100.0 (81.5-100.0) | 79/79, 100.0 (95.4-100.0) |
| Male | 26/30, 86.7 (69.3-96.2) | 50/53, 94.3 (84.3-98.8) |

| | | |
|---|---|---|
| Persons living with HIV | 7/9, 77.8 (40.0-97.2) | 23/25, 92.0 (74.0-99.0) |
| Persons living with diabetes | 10/11, 90.9 (58.7-99.8) | 19/21, 90.5 (69.6-98.8) |

**Table S4. Diagnostic accuracy of sputum swabs tested with MiniDock MTB compared to MRS by subgroups.**

| | Sensitivity n/N, % (95% CI) | Specificity n/N (95% CI) |
|---|---|---|
| Overall | 62/69, 89.9 (80.2-95.8) | 219/223, 98.2 (95.5-99.5) |
| Country | | |
| Uganda | 58/62, 93.5 (84.3-98.2) | 164/168, 97.6 (94.0-99.3) |
| Vietnam | 4/7, 57.1 (18.4-90.1) | 55/55, 100.0 (93.5-100.0) |
| Sex | | |
| Female | 20/24, 83.3 (62.6-95.3) | 121/121, 100.0 (97.0-100.0) |
| Male | 42/45, 93.3 (81.7-98.6) | 98/102, 96.1 (90.3-98.9) |
| Persons living with HIV | 8/11, 72.7 (39.0-94.0) | 44/46, 95.7 (85.2-99.5) |
| Persons living with diabetes | 17/18, 94.4 (72.7-99.9) | 35/37, 94.6 (81.8-99.3) |

**Table S5. Overall diagnostic accuracy of MiniDock MTB and comparator tests against a primary microbiological reference standard (MRS), combined test result.**

| | Sensitivity n/N, % (95% CI) | Specificity n/N (95% CI) |
|---|---|---|
| Tongue swab MiniDock MTB | 61/71, 85.9 (75.6,93.0) | 226/226, 100.0 (98.4,100.0) |
| Sputum swab MiniDock MTB | 66/70, 94.3 (86.0, 98.4) | 218/222, 98.2 (95.5,99.5) |
| Sputum Xpert Ultra | 67/71, 94.4 (86.2, 98.4) | 224/226, 99.1 (96.8, 99.9) |
| Sputum smear microscopy | 47/71, 66.2 (54.0, 77.0) | 226/226, 100.0 (98.4,100.0) |

**Table S6. Concordance of sputum swabs tested with MTB Ultima and sputum Xpert Ultra semi-quantitative grade.**

| | Positive concordance n/N, % (95% CI) | Negative concordance n/N, % (95% CI) |
|---|---|---|
| Overall (excluding Trace) | 45/49, 91.8 (80.4-97.7) | 142/143, 99.3% (96.2-100.0) |
| Overall (including Trace) | 46/54, 85.2 (72.9, 93.4) | 142/143, 99.3% (96.2, 100.0) |
| Semi-quantitative grade | | |
| Trace | 1/5, 20.0 (0.5, 71.6) | - |
| Very low | 2/3, 66.7 (9.4, 99.2) | - |
| Low | 12/15, 80.0 (51.9, 95.7) | - |
| Medium | 13/13, 100.0 (75.3, 100.0) | - |
| High | 18/18, 100.0 (81.5, 100.0) | |

**Table S7. Concordance of sputum swabs tested with MiniDock MTB and sputum Xpert Ultra semi-quantitative grade.**

| | Positive concordance n/N, % (95% CI) | Negative concordance n/N, % (95% CI) |
|---|---|---|
| Overall (excluding Trace) | 62/64, 96.9 (89.2-99.6) | 244/246, 99.2% (97.1-99.9) |
| Overall (including Trace) | 64/68, 94.1 (85.6-98.4) | 244/246, 99.2 (97.1, 99.9) |
| Semi-quantitative grade | | |
| Trace | 2/4, 50.0 (6.7, 93.2) | - |

| | | |
|---|---|---|
| Very low | 2/3, 66.7 (9.4, 99.1) | - |
| Low | 19/20, 95.0 (75.1, 99.8) | - |
| Medium | 17/17, 100.0 (80.5, 100.0) | - |
| High | 24/24, 100.0 (85.7, 100) | |

**Table S8. Diagnostic accuracy of tongue swabs tested with MTB Ultima compared to MRS by subgroups.**

| | Sensitivity n/N, % (95% CI) | Specificity n/N (95% CI) |
|---|---|---|
| Overall | 116/149, 77.9 (70.3-84.2) | 805/820, 98.2 (97.0-99.0) |
| Country | | |
| India | 26/35, 74.3 (56.7-87.5) | 363/374, 97.1 (94.8-98.5) |
| Uganda | 65/77, 84.4 (74.4-91.7) | 252/253, 99.6 (97.8-100.0) |
| Vietnam | 25/37, 67.6 (50.2-82.0) | 190/193, 98.4 (95.5-99.7) |
| Sex | | |
| Female | 38/51, 74.5 (60.4-85.7) | 390/392, 99.5 (98.2-99.9) |
| Male | 78/99, 78.8 (69.4-86.4) | 404/418, 96.7 (94.4-98.2) |
| Persons living with HIV | 12/16, 75.0 (47.6-92.7) | 85/85, 100.0 (95.8-100.0) |
| Persons living with diabetes | 37/41, 90.2 (76.9-97.3) | 183/187, 97.9 (94.6-99.4) |

**Table S9. Diagnostic accuracy of tongue swabs tested with MiniDock MTB compared to MRS by subgroups.**

| | Sensitivity n/N, % (95% CI) | Specificity n/N (95% CI) |
|---|---|---|
| Overall | 60/70, 85.7 (75.6-93.0) | 226/226, 100.0 (98.4-100.0) |
| Country | | |
| Uganda | 55/63, 87.3 (75.7-95.5) | 170/170, 100.0 (97.9-100.0) |
| Vietnam | 5/7, 71.4 (29.0-96.3) | 56/56, 100.0 (93.6-100.0) |
| Sex | | |
| Female | 19/24, 79.2 (57.8-92.9) | 121/121, 100.0 (97.0-100.0) |
| Male | 41/46, 89.1 (76.4-96.4) | 105/105, 100.0 (96.5-100.0) |
| Persons living with HIV | 8/11, 72.7 (39.0-94.0) | 46/46, 100.0 (92.3-100.0) |
| Persons living with diabetes | 16/18, 88.9 (65.3-98.6) | 39/39, 100.0 (91.0-100.0) |

**Table S10. Performance of index tests compared to MRS by induced/expectorated sputum.**

| | Sensitivity n/N, % (95% CI) | Specificity n/N, % (95% CI) |
|---|---|---|
| **MTB Ultima on tongue swab** | | |
| Expectorated | 106/135, 78.5 (70.6, 85.1) | 710/721, 98.5 (97.3, 99.2) |
| Induced | 10/14, 71.4 (41.9, 91.6) | 95/99, 96.0 (90.0, 98.9) |
| **MTB Ultima on sputum swab** | | |
| Expectorated | 1/3, 33.3 (0.8, 90.6) | 20/20, 100.0 (83.2,100.0) |
| Induced | 43/45, 95.6 (84.9, 99.5) | 109/112, 97.3 (92.4, 99.4) |
| **Pluslife MTB on tongue swab** | | |

| | | |
|---|---|---|
| Expectorated | 58/66, 87.9 (77.5, 94.6) | 202/202, 100.0 (98.2,100.0) |
| Induced | 2/4, 50.0 (6.8, 93.2) | 24/24, 100.0 (85.8, 100.0) |
| **Pluslife MTB on sputum swab** | | |
| Expectorated | 59/65, 90.8 (81.0, 96.5) | 195/199, 98.0 (94.9, 99.4) |
| Induced | 3/4, 75.0 (19.4, 99.4) | 24/24, 100.0 (85.8, 100.0) |

**Table S11. Overall diagnostic accuracy of MTB Ultima and sputum comparator tests against a primary microbiological reference standard (MRS), combined test result.**

| | Sensitivity n/N, % (95% CI) | Specificity n/N (95% CI) |
|---|---|---|
| **MTB Ultima tongue swabs and sputum-based comparator tests compared to MRS** | | |
| Tongue swab MTB Ultima | 116/150, 77.3 (69.8, 83.8) | 810/826, 98.1 (96.9, 98.9) |
| Sputum Xpert Ultra | 139/150, 92.7 (87.3, 96.3) | 814/826, 98.5 (97.5, 99.2) |
| Sputum smear microscopy | 88/150, 58.7 (50.3, 66.6) | 823/826, 99.6 (98.9, 99.9) |
| **MTB Ultima sputum swabs and sputum-based comparator tests compared to MRS** | | |
| Sputum swab MTB Ultima | 44/48, 91.7 (80.0, 97.7) | 129/132, 97.7 (93.5, 99.5) |
| Sputum Xpert Ultra | 48/48, 100.0 (92.6, 100.0) | 129/132, 97.7 (93.5, 99.5) |
| Sputum smear microscopy | 35/48, 72.9 (58.2, 84.7) | 132/132, 100.0 (97.2, 100.0) |

**Table S12. Concordance of tongue swabs tested with MTB Ultima and sputum Xpert Ultra semi-quantitative grade.**

| | Positive concordance n/N, % (95% CI) | Negative concordance n/N, % (95% CI) |
|---|---|---|
| Overall (excluding Trace) | 114/136, 83.8 (76.5-89.6) | 861/872, 98.7% (97.8-99.4) |
| Overall (including Trace) | 120/160, 75.0 (67.7, 81.5) | 861/872, 98.7% (97.8, 99.4) |
| Semi-quantitative grade | | |
| Trace | 6/24, 25.0 (9.7, 46.7) | - |
| Very low | 6/14, 42.8 (17.6, 71.1) | - |
| Low | 31/43, 72.1 (56.3, 84.6) | - |
| Medium | 31/33, 93.9 (79.8, 99.3) | - |
| High | 46/46, 100.0 (92.3, 100) | |

**Table S13. Concordance of tongue swabs tested with MiniDock MTB sputum Xpert Ultra semi-quantitative grade.**

| | Positive concordance n/N, % (95% CI) | Negative concordance n/N, % (95% CI) |
|---|---|---|
| Overall (excluding Trace) | 59/65, 90.8 (81.0-96.5) | 249/249, 100.0 (98.5-100.0) |
| Overall (including Trace) | 60/70, 85.7 (75.3, 92.9) | 249/249, 100.0 (98.5, 100.0) |
| Semi-quantitative grade | | |
| Trace | 1/5, 20.0 (0.5, 71.6) | - |
| Very low | 2/3, 66.7 (9.4, 99.2) | - |
| Low | 12/15, 80.0 (51.9, 95.7) | - |
| Medium | 13/13, 100.0 (75.3, 100.0) | - |
| High | 18/18, 100.0 (81.5, 100.0) | |
